# Association between extreme temperature events and dengue risks in Dhaka City, Bangladesh

**DOI:** 10.64898/2026.03.02.26347403

**Authors:** Ahnaf Shahriyar, S.M. Manzoor Ahmed Hanifi, Sheikh Mokhlesur Rahman

## Abstract

**Background:** Dengue outbreaks have become a severe threat to Bangladesh as the infections and mortality numbers are skyrocketing in recent years. Favorable environmental and anthropogenic conditions have established the capital of Bangladesh, Dhaka city as the epicenter of dengue outbreak. Studies have showed that climate change induced extreme weather events are exacerbating Aedes mosquito breeding and dengue virus transmission conditions.

**Methodology/Principal Findings:** In this study, short-term (0-6 weeks) associations of maximum temperature and heatwave days on dengue cases in Dhaka city were examined through Distributed Lag Non-linear Model (DLNM) methodology for weekly measurement of 2016-2024, taking into account relative humidity, cumulative rainfall, seasonality and hospital closure effect. Two separate negative binomial models were constructed. The maximum temperature model rendered an overall inverted U-shaped association, where the maximum temperature range of 31.5-33.2°C showed a sustained elevated dengue risk, with highest risk estimate at 33.2°C [relative risk (RR): 1.186, 95% CI: 1.002, 1.403]. Whereas, results of weekly heatwave days showed an overall protective effect (RR<1) for dengue cases. The lowest risk of infection was found at 3 heatwave days per week, with RR 0.275 (95% CI: 0.178, 0.423). Multiple sensitivity analyses were conducted for both models to evaluate their robustness. Lastly, the optimized models were analyzed under three distinct sub-periods, to capture the association of exposure variables with predominant circulating serotypes.

**Conclusions/Significance:** The findings of the study aim to support public health policymakers and healthcare authorities in designing and implementing effective vector control interventions under emerging climatic emergencies.

**Author Summary:** Dengue disease is one of the most buringing issue in Bangladesh in recent years. This vector-borne disease is inherently influenced by climatic variables, i.e., temperature, rainfall, humidity, etc. Moreover, these relations are complex and non-linearly associated. Due to shift in climatic conditions, the occurance of extreme weather events are becoming frequent, with increased magnitude and longer duration. In this study, the nonlinear and delayed association of dengue infections due to the exposure of extreme temperature events were assessed in climate-change vulnerable Dhaka city. To do this, a statistical method was used, called distributed lag nonlinear methodology (DLNM). The results showed that dengue infections had an inverted U-shaped (parabolic) relationship with maximum temperature, while compared to mean maximum temperature, and a suppressive association with heatwaves relative to days without heatwaves. The findings aim to work as an early warning system, and support to policymakes and healthcare authorities to tackle the dengue surge in the changing climate.

## Introduction

Dengue infection is a major public health concern in Bangladesh. The reported dengue fever cases in Bangladesh have increased more than eighteen-fold in the span of twenty-five years (from 5,500 cases in 2000 to 101,200 cases in 2024), with the capital Dhaka city being the epicenter of all major outbreaks [1,2]. Apart from high population density, the prevalence of dengue infections in Dhaka city is significantly influenced by favorable climatic factors, such as excessive rainfall, temperature, and humidity [1,3]. Moreover, climate change is likely to accelerate dengue spread at a local-level through rising temperatures and extreme weather events such as heat waves and droughts, as mosquito vectors can adapt to environmental and climatic changes [4–6]. The evolving relationship between dengue, human populations, and the environment suggests that climate change will shift dengue risk by altering mosquito biology, particularly in rapidly urbanizing tropical and subtropical regions [6].

Climatic factors in general are heavily linked to the biology of *Aedes* mosquitoes and transmission of dengue virus (DENV) infections. Mosquitoes are ectothermic organisms, meaning that environmental temperature influences nearly every stage of their lifecycle [7]. Temperature affects human exposure to mosquitoes [8], pathogen incubation period [9], mosquito bite rate [10], and *Aedes* mosquito growth rate, larval development and gonotrophic cycle duration [11,12]. Studies have further showcased that elevated temperatures within a certain range increase the *Aedes* mosquito replication rate and the risk of dengue transmission [13,14]. Conversely, virus replication and mosquito populations have been documented to decline at extreme temperatures [15,16]. Prior studies found that sustained extreme temperatures, i.e., heatwaves, exert an overall inhibitive effect on mosquito abundance [17,18]. Additionally, heatwave exposure demonstrated lower dengue infection risk at short-term lags (0-6 weeks), though statistically significant elevated risk was observed at extended lag periods (>7 weeks) [19]. Thus, the associations between temperature, heatwaves, and dengue infections are complex and exhibit contrasting non-linear effects with temporal variation [20].

Given that relationships between weather variability and health outcomes are typically non-linear and involve lagged effects, a simple linear association between exposure and response variable can lead to biased risk estimates and misleading interpretations [21]. Moreover, extensions of linear distributed lag models (DLMs) are insufficient in capturing these complex linkages, as they assume linear dose-response relationships and cannot capture complex bi-dimensional dependencies between predictor intensity and lag. Armstrong [22] and Gasparrinia et al. [23] addressed this shortcoming through introducing advanced statistical methods, such as the Distributed Lag Non-linear Models (DLNMs). In the DLNM framework, non-linear and lagged effects of exposure are simultaneously captured through a “*cross-basis*” function, which defines a bi-dimensional matrix across the predictor values and their corresponding lag periods [23]. This methodology has previously been extensively employed to investigate environmental variability and its non-linear lagged effects on response variables, such as dengue incidence worldwide [24–26].

While previous studies have assessed time-lagged nonlinear associations between temperature, heatwaves, and dengue infections across tropical and subtropical countries, Bangladesh-specific research remains necessary, as appropriate lag structures and association patterns are context-dependent on disease and location [27]. Prior studies in Bangladesh employed multiple linear regression and multivariate generalized linear regression models and found significant positive associations between temperature and reported dengue incidence [28–30]. However, these studies primarily focused on the linear effects of temperature on dengue and considered fixed lag structures, rather than distributed lags. Sharmin et al. [31] previously assessed the nonlinear association of reported monthly dengue cases in Bangladesh’s Dhaka district (2000-2009) with monthly diurnal temperature range and rainfall, using a negative binomial generalized linear model and adopting a Bayesian approach, while Hossain [28] employed quasi-Poisson and zero-infated Poisson regression model to evaluate the nonlinear relationship between climate variables (monthly total rainfall and daily average temperature) and dengue fever cases in Bangladesh from 2000 to 2021. Although in these studies, the lagged effects of the climatic variables were not assessed. Previously, in another study, Banu et al. examined the simultaneous nonlinear and lagged effects of ENSO and Indian Ocean Dipole (IOD) on dengue incidence in Bangladesh, adjusting for temperature and humidity, using the monthly dengue incidence rate from 2000 to 2012 [32]. However, apart from only an investigation by Banu et al. based on monthly measurements taken between 2000 and 2010, studies on the simultaneous non-linear and distributed-lagged association of maximum temperature on dengue cases in Bangladesh at finer temporal resolution remain sparse [33]. Similarly, the effects of heatwaves on dengue incidence research are rare, limited to descriptive and remote-sensing assessments of dengue clustering in urban heat island zones [34,35]. Thus, there remains a substantial research gap in understanding short-term, time-delayed exposure-response associations between heatwaves and dengue infections in the Bangladesh context.

The aim of the study is to examine the non-linear association between maximum ambient temperature and heatwaves on dengue cases in the outbreak epicenter of Bangladesh, Dhaka city, across appropriate lag structures, using historical data. It is anticipated that Bangladesh will experience rising temperatures and more frequent, intense heatwaves in the future [5]. Thus, investigating the association of dengue endemicity with the emerging climatic vulnerabilities has become a paramount concern. Gaining insight into these relationships could guide policymakers to design an effective early warning system, prevention strategies and optimal timing of implementation in the forthcoming extreme climatic scenarios.

## Methods

### Study area

Dhaka city, the capital of Bangladesh and a major megacity, is home to approximately 10.28 million people [36]. The city has been regarded as the dengue epicenter in Bangladesh. Moreover, Bangladesh’s dengue surveillance system is mostly Dhaka city-based [37].

### Data

#### Dengue data

Reported daily dengue cases for Dhaka city were collected from the Directorate General of Health Services (DGHS), Bangladesh, from January 2014 to September 2024 [2]. DGHS receives reports of suspected, probable, and confirmed dengue cases admitted or treated at various government and private healthcare facilities in Dhaka city [28,29]. Due to significant gaps in data from 2014 to 2015, the study focused on data collected between 2016 and 2024. Weekly counts of dengue cases were calculated by aggregating daily case reports across each week.

#### Meteorological data

Daily data on average temperatures (maximum, average), total rainfall and average relative humidity were obtained from the Bangladesh Meteorological Department (BMD) for Dhaka city’s meteorological station for the same timeframe. Absolute humidity (g/m^3^) was calculated using mean temperature and relative humidity values [Supporting Information (SI), S1 Eq.] [18]. For weekly data, daily climate readings were aggregated by week, and arithmetic means were computed to derive weekly averages of ambient temperature (mean and maximum), relative humidity and absolute humidity. Weekly cumulative rainfall was calculated as the sum of daily values over a week.

#### Hospital closure days

To account for the number of hospital closure days in Bangladesh due to public holidays and the medical holiday (Friday), a categorical variable was constructed. For weekly analysis, the total count of holidays within each week was aggregated. This variable was included in the model to adjust for the immediate effects of hospital shutdown due to public and medical holidays.

#### Population data

The annual population data of Dhaka city for 2015-2024 was collected from the World Bank Group Data Portal (population in the largest city, Bangladesh) [38]. This mid-year annual census data of Dhaka city was used to interpolate weekly and daily population estimates.

### Data analysis and model development

#### Estimation of heatwave days

A universally accepted definition of a heatwave day has not yet been established. The Bangladesh Meteorological Department (BMD) classifies a heatwave as an event in which the temperature exceeds 36℃ over a sizable area and persists for at least three days [39]. Nissan et al. recommended using a definition based on daily maximum and minimum temperatures exceeding the 95^th^ percentile for three consecutive days for Bangladesh [40]. In this study, a conservative method for defining heatwaves was adopted, aligning with a previous study of Seah et al., which defined heatwave days as periods when maximum temperatures surpassed the 90^th^ percentile value (35.5℃) for two consecutive days (S1 Table) [18]. The total number of such heatwave days within each epidemiological week (*t*) was summed to construct a continuous variable. This definition allows for the inclusion of heatwaves that extend across two consecutive epidemiological weeks. To minimize the risk of statistically significant findings arising from a single heatwave day definition, several alternative definitions were also evaluated. Details of the different heatwave definitions can be found in S1 Fig.

#### Distributed lag non-linear model (dlnm)

The exposure-lag-response of weekly maximum temperature (MaxT) and weekly heatwave days (HW) on reported dengue occurrences was examined using the Distributed Lag Non-Linear Model (DLNM), implemented through the R package ‘*dlnm*’ (version 2.4.10) [41]. The reported dengue cases were modelled using Generalized Linear Models (GLMs), specifically the negative binomial distribution. Two separate models were constructed using weekly estimates: (1) the MaxT Model and (2) the Heatwave Model. As the dengue transmission cycle is estimated to span 28 to 47 days (approximately 7 weeks) based on biological parameters, our models also accounted for climatological influences over a 7-week period, incorporating the immediate impact at lag 0 and delayed effects from weeks 1 to 6 [18].

In addition to the primary predictors (maximum temperature and heatwave days), both models included weekly mean relative humidity and weekly cumulative rainfall as potential confounders, along with two categorical variables (month of the year and weekly total hospital closure days). To address serial autocorrelation in both models, Autocorrelation Function (ACF) and Partial Autocorrelation Function (PACF) plots of the penultimate models were analyzed, and lags of the deviance residuals (*res*_*l*_) were incorporated based on the extent of residual autocorrelation detected. Additionally, the Durbin-Watson test was performed, and the *res*_*l*_ configuration yielding a statistic closest to 2 was selected to minimize

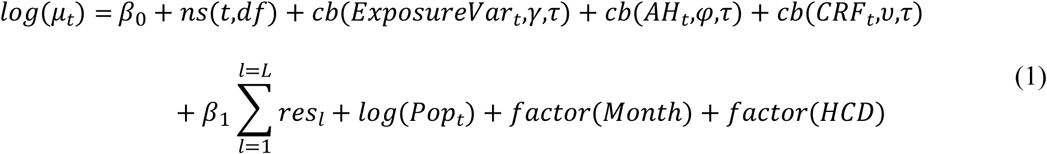

Where, *log*(μ_*t*_) is the expected number of weekly dengue reports on a week t with a log form and *β*_0_ is the intercept. The long-term trend and seasonality are adjusted using natural cubic splines, *ns* (*t*,*df*) with 3 degrees of freedom (DOF) per year. The c*b*(*ExposureVar*_*t*_,*γ*,*τ*) represent the cross-basis function of the exposure variable, weekly maximum temperature for the MaxT model and total number of heatwave days in week *t* for the HW Model. *cb*(*AH*_*t*_,*φ*,*τ*) and *cb*(*CRF*_*t*_,𝜐,*τ*) denote the weekly average relative humidity (*AH*_*t*_), and the weekly cumulative total rainfall (*CRF*_*t*_), respectively. *τ* denotes the number of lagged weeks (up to *τ* = 6-weeks lag). Furthermore, *γ*, *φ* and 𝜐 describe the coefficient vectors for the *ExposureVar*_*t*_, *AH*_*t*_ and *CRF*_*t*_ respectively. The non-linear and lagged effects of the cross-basis matrix were modeled using a natural spline (ns) smoothing function with 3 DOF for both the exposure and lag dimension [18,26,42]. *β*_1_ represents the coefficient vector for *res*_*l*_ and the population offset term, *log*(*Pop*_*t*_), reflects the logarithm of weekly population estimates in Dhaka city. Finally, the models also incorporated two separate categorical independent variables, month of the year (*Month*) and hospital closure days in Bangladesh (*HCD*), denoted by ‘*factor*(*Variable*)’.

#### Collinearity and model performance assessment

Collinearity between independent variables in the final models specified in Equation (1 was evaluated using the Generalized Variance Inflation Factor (GVIF) [43]. A threshold value of 2 was used to indicate potential multicollinearity [18]. To evaluate model performance, likelihood ratio tests were employed and the full model was compared against a null model containing only an intercept [21]. Lastly, sequential deviance analyses (Type I ANOVA) were then used to examine how each predictor set incrementally improved model fit.

#### Sensitivity analysis

Sensitivity analyses were conducted to evaluate the robustness of model estimates to different specifications. In the MaxT model, spline functions of time were tested with varying DOF per year to capture trend and seasonal patterns. Sensitivity analyses were further performed by varying the degrees of flexibility used to model the non-linear and lagged effects in the MaxT cross-basis. For the HW model, three sets of sensitivity analyses were conducted. First, the degree of control for long-term trend and seasonality was varied by changing the spline DOF per year. Second, alternative specifications of the heatwave cross-basis function were tested using different internal knot placements. Lastly, alternative definitions of heatwave days were used to evaluate the consistency of the observed associations.

#### Measure of effect

Results from both models were reported as relative risk (RR) [44], with corresponding 95% confidence intervals (CI). The mean values were used as the baseline reference value for calculating RR for each weather parameter [18,33]. However, for heatwave days, the reference value was defined as no heatwave days in the week (HW = 0). All statistical tests were two-tailed, with p-values less than 0.05 considered statistically significant.

#### Sub-period analysis

All four DENV serotypes (DENV-1 to DENV-4) are prevalent in Dhaka, though the dominant circulating serotype varied with time [45,46]. For instance, from 2016 to 2018, the DENV-2 serotype was prevalent, while the DENV-3 serotype dominated from 2019 to 2022 [45]. Beyond 2022, DENV-2 once again became the predominant serotype [47]. Hence, to capture the association of dominant dengue serotype with the exposure variables, the final optimized MaxT and HW model was further analyzed by dividing the study period into three Sub-periods: Sub-period 1 (2016-2018, DENV-2), Sub-period 2 (2019-2022, DENV-3), and Sub-period 3 (2023-2024, DENV-2).

## Results

### Descriptive statistics

Bangladesh experienced the most severe dengue incidences in 2023 and 2019, reporting a total of 380,492 and 113,090 cases, respectively, of which Dhaka city accounted for 28.9% of cases in 2023 and 45.8% in 2019 (S2 Fig). Among all the regions, Dhaka city showed clear peaks of dengue cases throughout the reported years. Additionally, ‘large outbreaks’ have been occurring in Dhaka city from 2019 onwards (S3 Fig), except in 2020, which may be attributed to the disruptions in dengue surveillance due to the COVID-19 pandemic [1]. In terms of seasonality, peak dengue transmissions in Dhaka city typically occur during the monsoon months (June-September), while winter and pre-monsoon months (December-April) consistently show minimal transmission throughout the study period (Table 1). S4 Fig illustrates the time series plot of daily climatic parameters of Dhaka city from 2016 to 2024. All the parameters, including dengue cases, show seasonal patterns and are thus time-dependent.

**Table 1:** Number of dengue patients in Dhaka city by year and month.

|  | Jan | Feb | Mar | Apr | May | Jun | Jul | Aug | Sep | Oct | Nov | Dec |
| --- | --- | --- | --- | --- | --- | --- | --- | --- | --- | --- | --- | --- |
| 2016 | 14 | 3 | 15 | 42 | 66 | 246 | 824 | 1423 | 1460 | 1020 | 522 | 141 |
| **2017** | 82 | 57 | 21 | 63 | 116 | 252 | 285 | 334 | 415 | 509 | 394 | 125 |
| **2018** | 25 | 6 | 19 | 29 | 52 | 295 | 946 | 1795 | 3086 | 2405 | 1192 | 293 |
| **2019** | 38 | 18 | 17 | 58 | 193 | 1865 | 13336 | 25161 | 6354 | 2514 | 1567 | 682 |
| **2020** | 147 | 35 | 26 | 23 | 8 | 18 | 20 | 66 | 43 | 154 | 493 | 191 |
| **2021** | 19 | 7 | 12 | 2 | 41 | 262 | 2241 | 6955 | 6343 | 4357 | 2790 | 586 |
| **2022** | 59 | 10 | 11 | 19 | 161 | 701 | 1283 | 2847 | 7190 | 13735 | 10491 | 2713 |
| **2023** | 272 | 76 | 74 | 92 | 864 | 4701 | 23121 | 28821 | 25201 | 15947 | 8667 | 2172 |
| **2024** | 358 | 123 | 119 | 162 | 221 | 328 | 1509 | 3038 | 7993 | - | - | - |

Spearman correlation analysis was performed with all the variables (S5 Fig), and strongly correlated variables (mean temperature, absolute humidity) were excluded from further analysis. Table 2 presents the descriptive statistics of the selected variables in the study. The study covered 456 epidemiological weeks, during which the weekly total dengue cases exhibited substantial variation and overdispersion, with a mean (SD) of 563.7 (1311.3). Moreover, the median number of dengue cases (59) is considerably lower than the mean, and the maximum value (7,987) is extremely high, indicating a right-skewed distribution.

**Table 2:**
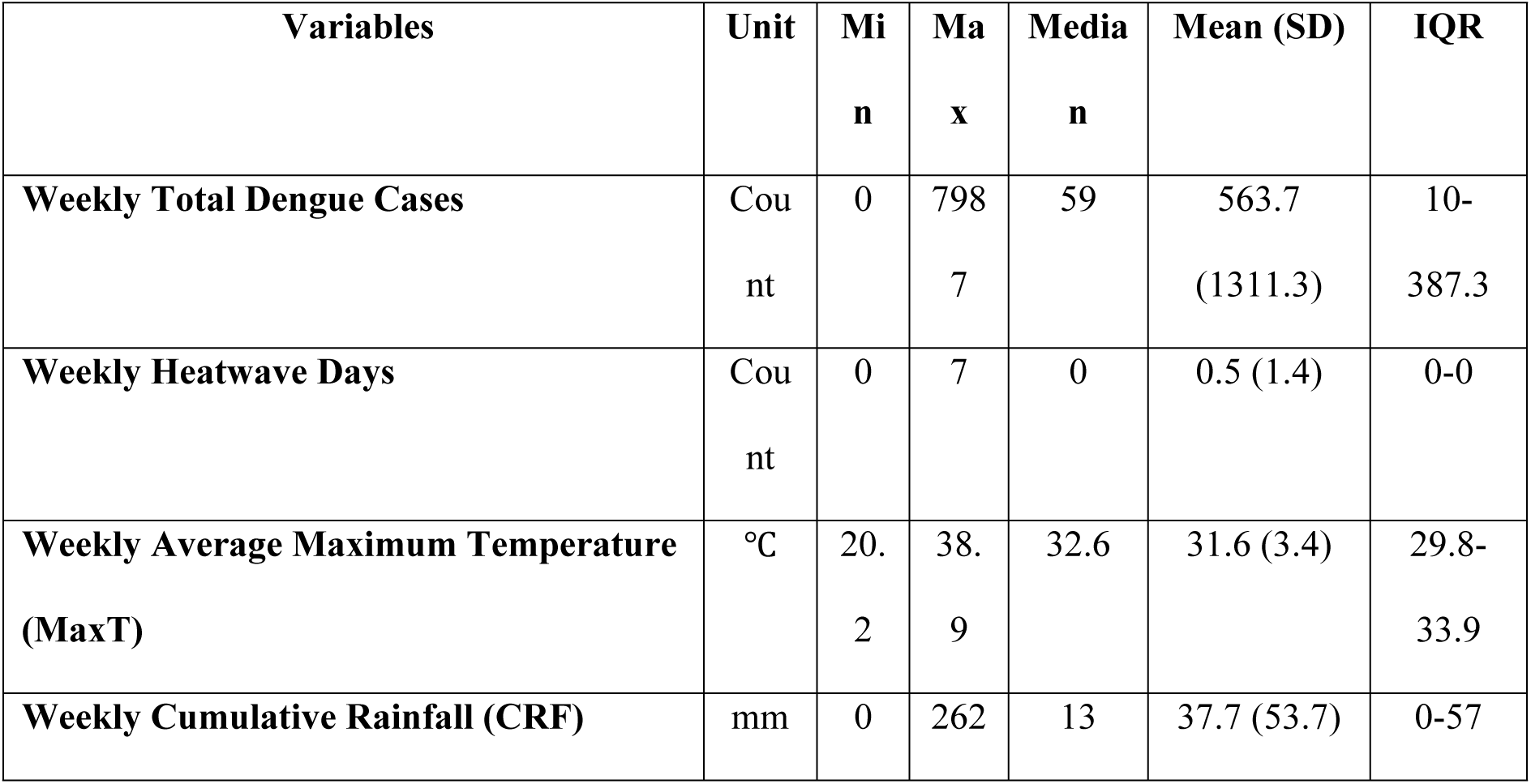
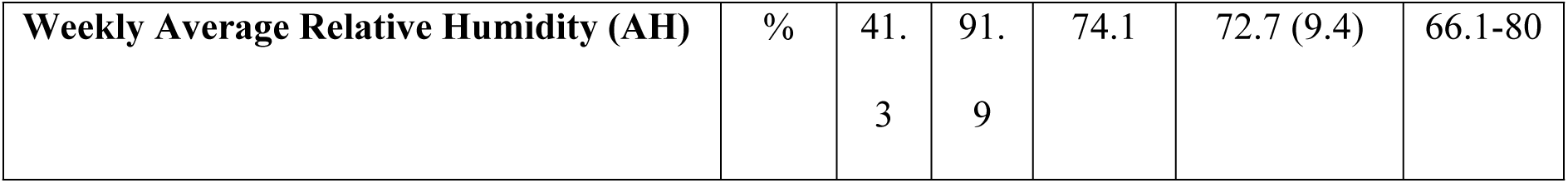
Summary statistics of weekly measures of dengue cases and meteorological variables in Dhaka City (2016-2024). Interquartile range (IQR) refers to the 1^st^ and 3^rd^ quartiles.

On the other hand, the zero median value of weekly heatwave days denotes that most weeks had no heatwave. The total number of heatwave days within a year was observed to be highest in 2024 with 51 days, followed by 2023 and 2014 with 49 days. Whereas 2018 had the lowest number of heatwave days (9 heatwave days). Heatwaves sustaining two days were most common (25 weeks), followed by one-day heatwaves (16 weeks), while five-day heatwaves were rare (S2 Table). Heatwaves that started on the last day of the previous epidemiological week and continued into the following week are referred to as one-day heatwave days. It is evident that changing the definition of heatwave alters the total count of week numbers with different heatwave days. Intuitively, increasing the threshold for the definition of heatwave days resulted in fewer heatwave day events. Thus, some extreme heatwave day definitions were excluded from further analysis.

### MaxT model

#### Model development

The collinearity, autocorrelation of the variables and significance of the whole model and individual terms of the MaxT model were evaluated. Multicollinearity assessment showed that all variables in the model had GVIF values below 2, indicating no significant collinearity among the predictors (S3 Table). The ACF and PACF plots indicated that some autocorrelation remained in the final adjusted MaxT model, though it was not strong (S6 Fig). The residual diagnostic plots indicate a reasonable goodness of fit. The MaxT model significantly outperformed the null model (likelihood ratio test: *χ*^2^ = 1812, df = 77, P < 0.001). Sequential deviance analyses revealed that all predictors contributed significantly to model fit (S4 Table).

#### Exposure-lag-response association

Fig 1(a) displays a 3D exposure-lag-response surface for maximum temperature and confirmed dengue cases risk, with RR estimated against a reference mean temperature of 31.5°C. The plot exhibited a very strong and immediate effect of the lowest maximum temperature (20°C). The RR remained above 1.00 up to approximately 32°C, after which it declined significantly, reaching its lowest point at the highest temperature (39°C) at lag 0. At longer lags, the plot suggested a harvesting pattern (morbidity displacement) following exposure to low maximum temperatures. In contrast, the temperature range of 31°C to 35°C was associated with a delayed but sustained elevated dengue risk, with RR consistently above 1.00 from lag week 1 through week 5, with risk attenuation at lag week 6. Lastly, the RR for maximum temperature greater than 35°C consistently remained below 1.00 throughout the entire lag structure, indicating a protective effect across all lag structures. To provide a more detailed assessment of the relationship, along with the associated uncertainty in the estimates, the exposure-response associations for each lag were plotted (S7 Fig).

**Fig 1.**
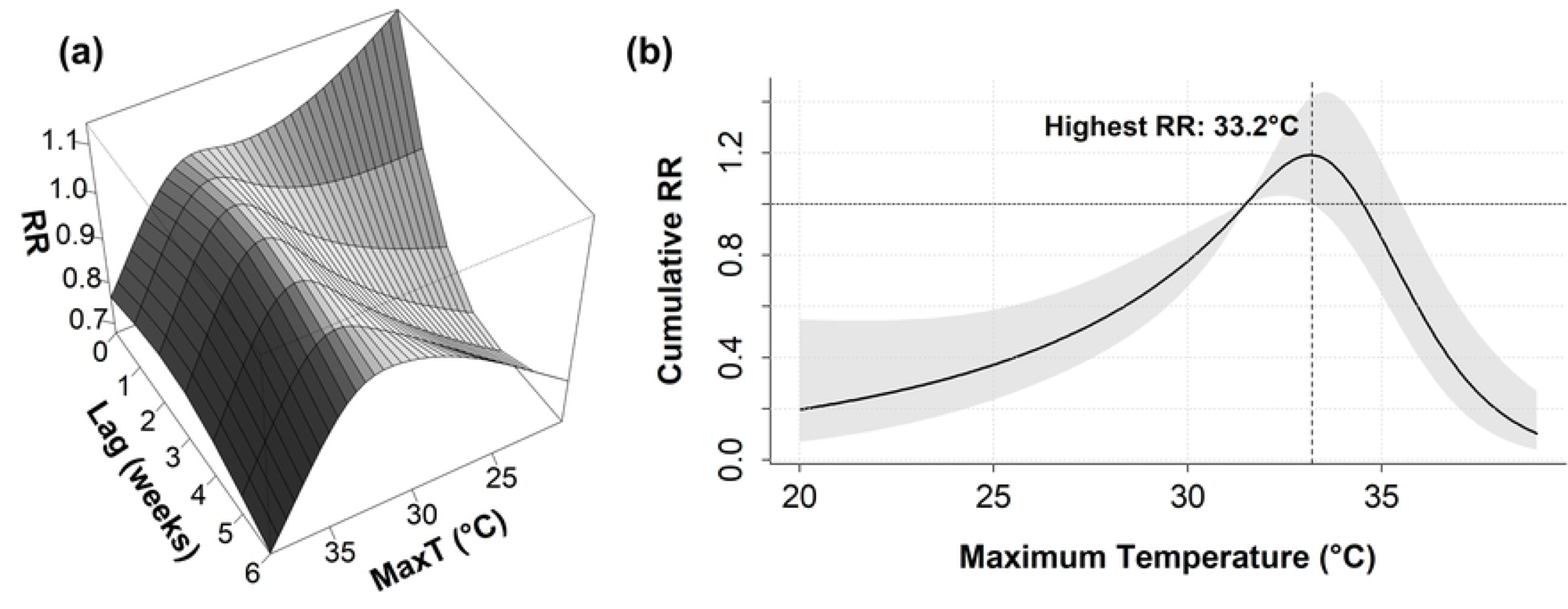
Exposure-response Relationship of the MaxT Model. (a) 3-D plot of RR along Maximum Temperature (MaxT) and Lags (Weeks), with reference at 31.5°C. (b) Cumulative exposure-response curve for MaxT and dengue RR over a 0-6 week lag, with mean MaxT (31.5°C) as the reference value. Shaded regions denote 95% confidence intervals.

#### Overall cumulative risk pattern

The overall cumulative exposure-response curve, relative to 31.5°C, revealed an inverted U-shaped (parabolic) association between MaxT and reported dengue infections (Fig 1b**Error! Reference source not found.**). The risk of dengue cases reduced steadily as the maximum temperature fell below the reference level (31.5°C). Beyond the base level, the risk of infections increased and peaked at 33.2°C with RR 1.186 (95% CI: 1.002, 1.403), which was 3% higher than the median maximum temperature. From 31.5-33.2°C MaxT range, statistically significant elevated dengue risk was observed. However, as the confidence boundary contains the value 1.00, the risk estimations of 33.3-35.5°C are not statistically significant [48]. The risk estimation declined back to baseline (RR: 1.00) at 34.5°C, and from this point onward, the risk of dengue cases decreased steadily, with the lowest risk observed at the highest maximum temperature of 38°C (RR: 0.191, 95% CI: 0.091, 0.403). Overall, a 1°C decrease from the mean maximum temperature was associated with a 16.1% reduction in dengue risk (RR: 0.839, 95% CI: 0.758, 0.928), while a 1°C increase corresponded to a 15% increase in risk (RR: 1.151, 95% CI: 1.031, 1.286).

#### Sensitivity analysis

Results of the sensitivity analysis using varying the degree of control for long term trend and seasonality in the MaxT Model are shown in S8 Fig. The direction of effect or the strength of association did not alter to a large degree from the original dose-response curve with the varying spline function DOF per year. In 5 DOF per year MaxT model, peak RR occurred in 33.9°C (RR: 1.370, 95% CI: 1.054, 1.781), and 1°C decrease and increase from the mean maximum temperature resulted in 20.5% reduction of risk (RR: 0.795, 95% CI: 0.715, 0.885) and 22.3% increase in risk (RR: 1.223, 95% CI: 1.087, 1.376), respectively. Similarly, in the 7 DOF per year MaxT model, peak RR occurred in 33.9°C (RR: 1.614, 95% CI: 1.202, 2.167). Risk estimates in this model showed a 24.5% reduction (RR: 0.755; 95% CI: 0.662, 0.861) and 33.1% increase in risk (RR: 1.331; 95% CI: 1.156, 1.532) for a 1°C decrease and increase, respectively. These findings suggest that the results are robust to the initial assumptions used in model development.

Moreover, the results of the sensitivity analysis by increasing the DOF used to model the non-linear and lag effects of MaxT are shown in S5 Table. Increasing the DOF of exposure and lag dimension from 3 to 4 and 5 did not alter the shape of the exposure-response curve, the direction of effect, nor the strength of association to a large degree. The model using a MaxT cross-basis with 4 DOF estimated the peak RR at 32.7°C (RR: 1.196, 95% CI: 1.055, 1.355), while the 5 DOF model identified the peak at 32.9°C with a higher risk estimate (RR: 1.458, 95% CI: 1.082, 1.965). The results of this model corroborate the baseline model estimations. Thus, the results with the initial assumptions of model development can be considered robust.

### Heatwave (HW) model

#### Model development

Similar to the MaxT model, the HW model also undergone multiple evaluation criteria. The multicollinearity diagnosis showed that the GVIF value for each variable is less than 2 and thus no serious collinearity existed within the model (S9 Fig). The ACF and PACF plots in S9 Fig indicated that some autocorrelation between deviance residuals was present; however, they were not strong. The deviance residuals of the developed model were nearly normally distributed and the plot of residuals vs log of observed dengue cases indicated a reasonably good fit. The HW model fit the data significantly better than the null model (χ^2^ = 1800, df = 80, p < 0.001). Sequential deviance results further indicated that each predictor contributed significantly to the overall model fit (S7 Table).

#### Exposure-lag-response association

Fig 2(a) represents a 3-D diagram of the risk estimation along weekly heatwave days and lag weeks compared to a reference value of zero heatwave days. The plot showed a very pronounced and immediate effect of the maximum number of heatwave days in a week (HW = 7). Risk estimates of 7 heatwave days reduced steeply at longer lag dimensions, suggesting a harvesting effect. The association further suggested protective effects at exposure to moderate heatwave days (2-4 days), especially at 0-4 week lag structures. Moreover, the exposure-response curve at lag week zero exhibited a non-monotonic, U-shaped pattern. The RR declined from the reference level (RR = 1.00) and reached its minimum at 3-4 weekly heatwave days. After this point, RR elevated, intersected the baseline at 6 heatwave days, and finally reached the highest RR estimation at weekly 7 heatwave days. This pronounced U-shaped association of RR and heatwave days was observed throughout lag week 0-1, then lag week 2-7 demonstrated a reverse J-shaped relationship, of which lag week 5-6 presented a rather unusual pattern with a declining tail at higher exposure frequencies. S10 Fig presents the exposure-response associations for each lag, providing a detailed assessment of the relationship, with the corresponding uncertainty in the estimates.

**Fig 2.**
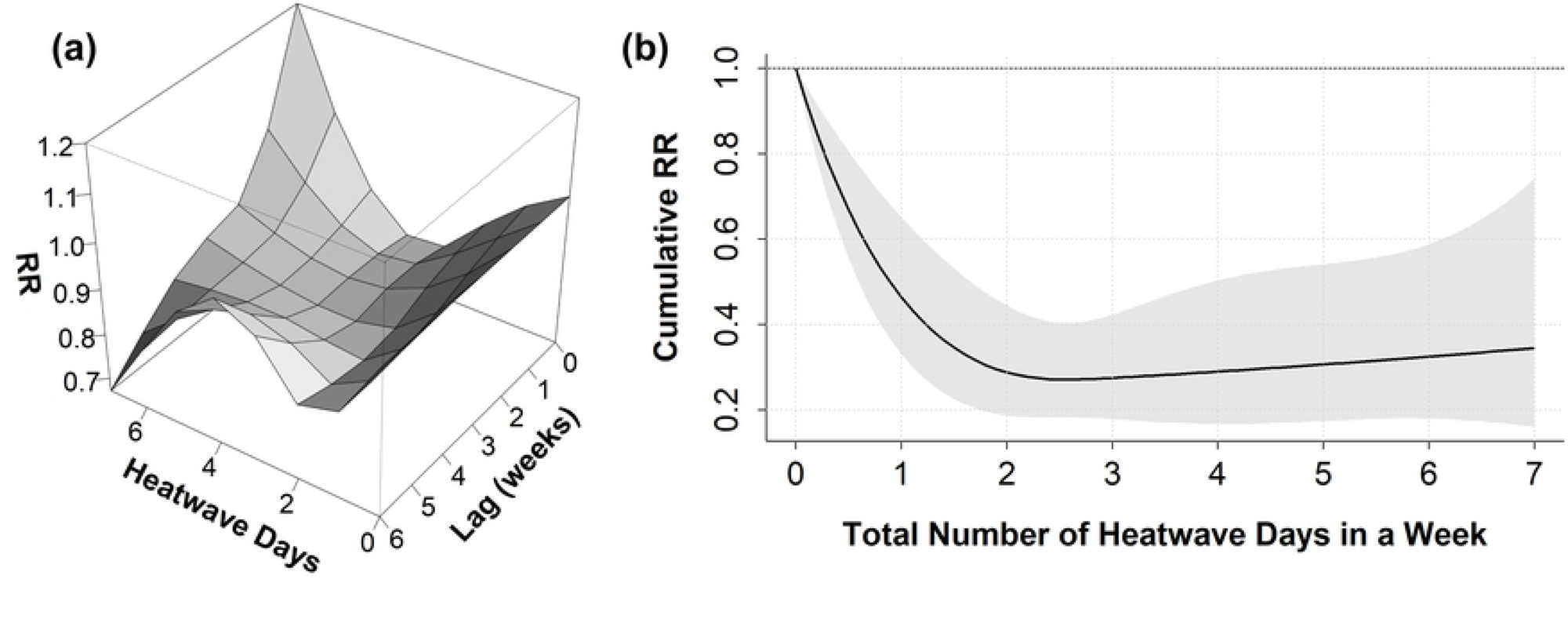
Exposure-response Relationship of the HW Model. (a) 3-D plot of RR along total Heatwave Days per week and Lags (Weeks), with reference to zero heatwave days. (b) Cumulative exposure-response curve for total Heatwave Days per week and dengue RR over a 0-6 week lag, with zero heatwave days as the reference value. Shaded regions denote 95% confidence intervals.

#### Cumulative risk association

Fig 2(b) illustrates the cumulative effect of the total number of weekly heatwave days on reported dengue infections, using zero heatwave days as the reference. A reverse J-shaped association with a strong protective effect from moderate to higher exposure levels was observed between the predictor and response variable. The risk of recorded dengue cases initially declined steeply from the reference level to 0.464 (95% CI: 0.331, 0.652) with exposure to 1 heatwave day. The highest protective effect is observed at three heatwave days per week with a 72.5% reduction (RR: 0.275, 95% CI: 0.178, 0.423), compared to the reference level. Beyond this point, the effect estimate showed an ever-so-slightly increasing trend with minimal variation across higher exposure levels. The CIs become progressively wider at higher exposure levels, particularly at 7 heatwave days (RR: 0.345, 95% CI: 0.160, 0.741), suggesting increased uncertainty in effect estimates at extreme exposure frequencies.

#### Sensitivity analysis

The outputs of the sensitivity analysis by varying the degree of control for long-term trend and seasonality in the Heatwave model are illustrated in S10 Fig. Varying the smoothness level of the spline function with 4, 5, and 7 DOF per year did not substantially alter the magnitude or direction of effect for 1-6 heatwave days. Overall, the 4 DOF per year model showed a U-shaped curve, with the highest risk reduction at moderate exposure levels and increased RR at 7 heatwave days from the baseline, which is suspected of overfitting. On the other hand, 5 and 7 DOF per year showed a similar association between the predictor and response variable as the base model. Across all models, the minimum RR values were consistently observed around 3-4 heatwave days per week. The outputs of this sensitivity analysis suggest that the initial results were robust.

Sensitivity analysis using different heatwave cross-basis specifications, as shown in S12 Fig, revealed that no notable changes in the direction or magnitude of the estimated effects occurred by varying the knot placements of the spline function in the heatwave cross-basis. At exposure to 4-6 heatwave days, some models rendered a hump-shaped pattern with a wide CI and were statistically non-significant, which can be attributed to the fewer counts of heatwave events at this frequency. At the maximum weekly heatwave day, the RR remained between 0.3 and 0.4 across all the models. Results under this sensitivity analysis further manifested the robustness of the initial model.

Additionally, the direction and strength of the association remained consistent for 1-3 heatwave days, regardless of the heatwave definition used in the sensitivity analysis (S13 Fig). Beyond the 3 heatwave days point, a hump-shaped association with large variability was observed for 4-6 heatwave days, similar to the previous sensitivity analysis. The model with maximum temperature exceeding the 80^th^ percentile for 3 consecutive days presented statistically significant increased risk in this frequency, while other models either corroborated the base-model or resulted statistically non-significant.

### Effects of climatic confounders on the models

#### Relative humidity

The overall cumulative effect of average relative humidity (AH) on RR of reported dengue infections for lag period 0-6 weeks is shown in S14 Fig, for both the Heatwave and MaxT model. The risk is calculated with reference to the mean value of AH (72.6%).

In the heatwave model, the risk of dengue cases was rendered statistically non-significant on both sides of the mean AH. The effect estimates due to AH in the MaxT model showed an increasing trend, starting from the lowest RR at the minimum AH and reaching the baseline (RR = 1.00) at 60% AH. The risk estimate then remained statistically non-significant at the reference value up to 80% AH, and finally, beyond 80% AH, it showed an increased risk estimate with wide CIs.

#### Cumulative rainfall

The overall cumulative effects of the weekly cumulative rainfall (with respect to the mean CRF) in the Heatwave days model and MaxT model are shown in S15 Fig. In the heatwave days model, the RR estimation increased steadily beyond the mean CRF value, however it was statistically non-significant. Similarly, effect estimates of weekly CRF on both sides of the mean value were statistically non-significant in the MaxT model.

### Exposure-response relationship in sub-periods

The overall effects of MaxT on dengue cases across a 0-6 week lag dimension in each sub-period are presented in Fig 3(a)-(c). In sub-period 1, a broadly linear relationship was observed between the maximum temperature and dengue cases, with the highest RR at the highest maximum temperature (statistically non-significant). While the remaining sub-periods showed the conventional parabolic relation with the exposure variable, with peak risk estimates at 34°C and 33.8°C, respectively.

**Fig 3.**
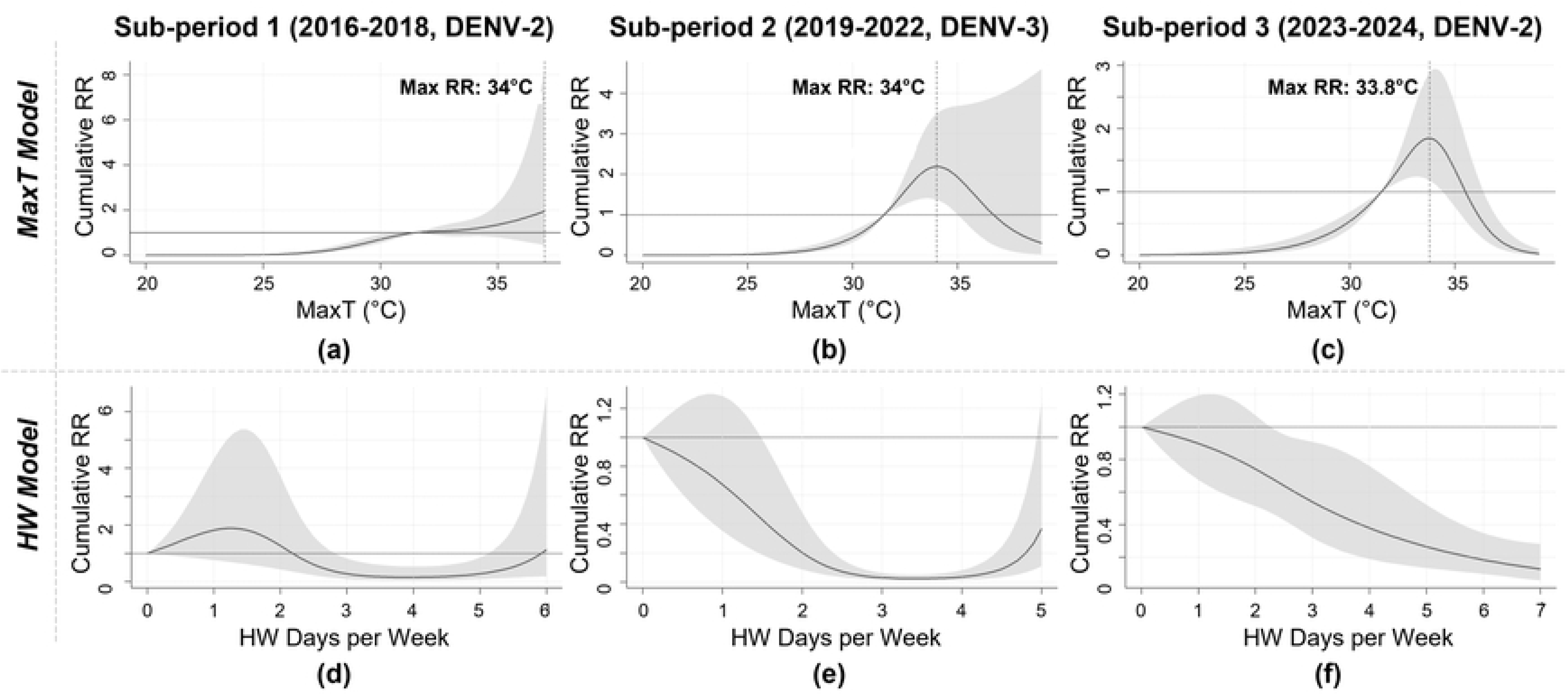
Cumulative exposure-response curve of dengue cases for each sub-period. (a)-(c): Effect of MaxT over a 0-6 week lag; (d)-(f): Effect of Heatwave days over a 0-6 week lag. The shaded region indicates 95% CI of the fitted RR. Reference MaxT = 31.5°C and HW = zero heatwave days.

Fig 3(d)-(f) displays the overall cumulative impact of heatwave days on dengue cases across each sub-period for the predefined lag period. The plots show RR estimates for the optimum number of weekly heatwave days, beyond which the extremely wide CIs render the plots illegible. Statistically significant reduced risk estimates were observed in 3-5 heatwave days for sub-period 1, weekly 2-4 heatwave days for sub-period 2, and 3-7 heatwave days for sub-period 3.

## Discussion

This quantitative study aimed to explore the non-linear and lagged association between maximum temperature and heatwave days on dengue cases in Bangladesh’s outbreak epicenter, Dhaka city, using historical data. In this quantitative study, the association between maximum temperature and dengue cases in Dhaka city followed a parabolic pattern with statistically significant elevated risk at a 32.7-33.9°C temperature range. While, heatwaves exerted a reverse J-shaped association with a short-term protective effect on dengue infections. Exposure to both of the primary predictor variables showed non-linear and lagged effects. These findings offer critical insights into the Dhaka city’s dengue dynamics under present and future climatic conditions on a broad level, as controlled laboratory settings to understand the biology of vectors under climatic stress are restricted by the quantity of subjects and time in laboratory experiments [17].

### Parabolic association with maximum temperature

Risk of dengue transmission has been documented to increase at elevated but not extreme temperatures [15,49]. In this study, the overall cumulative relationship between maximum temperature and dengue cases over a short-term (7-week) lag period was an inverted U-shaped (parabolic) curve. This association was similar to a study conducted in Singapore [18]. However, contrary to the Singapore research, this study found 31.5-33.2°C maximum temperature range to be the peak dengue risk estimate, instead of 31.1°C maximum temperature. Similar to a previous study in India by Kakarla et al., this study also found sustained elevated dengue risk within 31.5-34.5°C maximum temperature range throughout the shorter lag dimension [25]. Studies have found that the dominant strain responsible for the 2023 surge in Bangladesh (DENV type-2) exhibited threefold increased transmission rates at 32-35°C temperature range, compared to 30°C maximum temperature [49]. This is because the EIP reduces from 12 days at 30°C to just 7 days at higher temperatures. Prior research has also demonstrated that an elevation in ambient temperature within a specific range diminishes the virus’s EIP, lowers mortality rates, and enhances the *Aedes* mosquito’s blood-feeding behavior, biting rate and vectorial capacity [13,14,25]. As a result, mosquitoes may bite more frequently after feeding on an infected person, increasing the chances of transmission (possibly causing a fast surge) [50]. Moreover, at higher temperatures, infected adult vectors require more blood meal to complete the gonotrophic cycle, and multiple gonotrophic cycles throughout the vector’s lifespan increase transmission risk [15]. Arcari et al. illustrated that increasing temperatures can accelerate dengue fever transmission, often leading to more rapid and occasionally explosive outbreaks [51]. Furthermore, elevated ambient temperatures accelerate the *Ae. aegypti* life cycle, resulting in smaller mosquitoes [12]. Smaller females require more blood meals for egg development, increasing transmission and accelerating epidemic spread, even during the dry season. Additionally, increased dehydration due to higher temperatures may also enhance biting frequency through frequent human-mosquito contact, thereby intensifying transmission [12].

### Short-term protective effect of heatwaves

The overall cumulative dengue risk estimation over 0-6 week period was reverse J-shaped in the heatwave model of our study. Similar to a previous study in Singapore by Seah et al., our study also found a protective effect of dengue cases due to the exposure to 1-7 weekly heatwave days [18]. However, the results of our study exhibited a stronger protective effect compared to the Singapore study. At 3 heatwave days, approximately 44% less dengue risk estimate was observed in Dhaka city compared to Singapore. A study on *Ae. albopictus,* where heatwave is defined as daily maximum temperature greater than 35°C for three consecutive days, revealed that heatwave events tend to have an overall suppressive effect on dengue development [17]. Similarly, in Hanoi, Vietnam, at a short-term lag structure (0-6 weeks), risk of dengue infections was lower for all three types of outbreaks (small, medium and large) [19]. Although, at higher lag, all three outbreak types showed statistically significant increased risk: small outbreak at 7-week and 9-week, medium outbreak at 7-week and large outbreak at 14-week. In Guangzhou, China, the six-week delayed effects of the heatwave increased dengue outbreak risk by up to 251% [52]. The biological plausibility of this effect can be explained by the complex physiological stress responses in *Aedes* mosquitoes triggered by extreme heat. Zhao et al. [53], Sivan et al. [54] and Singh et al. [55] observed both downregulation and upregulation of several heat shock protein (HSPs) genes in *Aedes* mosquitoes when exposed to thermal stress (35-42°C) for 1-6 hours. Despite short-term acceleration of early life-stage development, sustained high temperatures produced an overall inhibitory effect on population growth in *Aedes albopictus* populations [17]. Mordecai et al. revealed that mosquito development, biting rate, fecundity, life span and transmission are found to be inhibited in an environment with extremely high temperatures (> 35°C) [56]. Moreover, one generational exposure to heat (sudden thermal fluctuation) resulted in fitness costs and increased resistance to viral infection in *Aedes* mosquitoes; whereas, warm-evolved mosquitoes did not suffer fitness costs and showed less resistance to viral infection [57].

### Association with climatic confounders

The rainy season is usually positively associated with infected adult dengue abundance and higher dengue transmission [58]. Kakarla et al. [25] found that dengue’s RR gradually rises from 40 to 60 mm of cumulative weekly rainfall, then falls when rainfall exceeds 80 mm, while findings of Benedum et al. [59] showed excess rainfall negatively affects dengue spread. Our study did not observe any statistically significant short-term association of dengue risk with rainfall, similar to previous studies [18,19,60]. These diverging relationships can be explained by the fact that, in urban and semi-urban locations, inadequate infrastructure and high water storage container density and waste volume enable year-round mosquito reproduction [25]. As a result, precipitation levels become a less direct determinant of dengue transmission in these settings.

In this study, significant association between average relative humidity and reported dengue infections was not observed for the majority of AH values. Similar findings were also found in a study in Bangladesh [61] and Singapore [60]. However, some studies observed significant positive associations between absolute humidity and risk of dengue cases [18,60], which was excluded from our analysis because it showed a strong correlation with rainfall and average humidity. AH value towards 90% showed an elevated risk estimate in our MaxT model, which aligns with the biology of *Aedes* mosquito [12].

### Association with predominant circulating serotype

During our whole study period, the serotype of DENV in Dhaka city shifted its circulating predominance multiple times. Serotype-specific immunity levels in the population change from one period to another [60]. Thus, it is important to assess the response of the predominant serotypes due to the effect of exposure variables during each time period. The association and peak risk estimation in DENV-2-dominated sub-period 1 differed from the remaining two sub-periods and the whole period in the MaxT model. The sub-period 1 showed a gradual increasing trend from the lowest MaxT value and the peak risk was observed at the highest maximum temperature, though it was non-significant. While, sub-period 2 (DENV-3 dominant) and sub-period 3 (DENV-2 prevalent) demonstrated peak dengue risk at 33.8-40°C, which was consistent with the whole period base model. In case of heatwave days, all the sub-periods showed statistically significant reduced risk of dengue cases at exposure to moderate heatwave days. Thus, the risk of dengue is inhibited due to heatwaves, regardless of the dominant serotype.

### Implications and limitations of the study

According to the IPCC, Bangladesh is predicted to experience a warming level of 1.5°C-2.0°C due to climate change, with frequent occurrences of extreme heatwaves [5]. Extreme temperatures and heatwaves will lead to a rise in vector-borne infectious diseases, which pose a significant risk of increased mortality and morbidity [5]. The consequences of this forecasted climate shift are already evident. In 2016, the mean maximum temperature of Dhaka city was 31.6°C with 24 heatwave days, which rose to 32.1°C with 51 heatwave days in 2024. Additionally, the number of reported dengue cases increased by 29,400 (more than 500%) over the same period. In this study, the statistically significant highest dengue risk for Dhaka city was observed at a maximum temperature range of 31.5-33.2°C, while sustained extreme temperatures (i.e., greater than 35.5°C) and heatwave events were found to lower the risk of dengue cases. These findings are crucial for dengue-devastated Dhaka city. If the maximum temperature of Dhaka shifts to this elevated range due to climate change, dengue cases would also exhibit an increase. However, this risk would be largely suppressed if the maximum temperature exceeded 35.5°C for a consecutive number of days.

This study aligns closely with the Strategic Objective 4 and Strategic Objective 6 of the Bangladesh National Dengue Prevention and Control Strategy (2024-2030) by directly addressing the Strategy’s emphasis on climate-sensitive surveillance, early warning, and evidence-based decision-making [62]. The findings of this study can be utilized to develop a web-based EWS, i.e., Dengue Risk Dashboard, in collaboration with the Directorate General of Health Services (DGHS), Institute of Epidemiology Disease Control and Research (IEDCR), icddr,b, Aspire to Innovate (a2i), etc. By providing real-time data, the system will aim to empower individuals to take proactive precautions at a personal level to mitigate their risk of dengue infection. Furthermore, it will act as a strategic tool for policymakers and healthcare authorities, guiding them in the development of data-driven prevention strategies and ensuring the optimal timing for the implementation of public health interventions.

There exists some limitations to the study. Out of several hundred hospitals and clinics across Dhaka city, the dengue surveillance system currently receives daily admission and fatality reports from only 77 hospitals (18 public and 59 private) [2]. Moreover, the current tracking system inherently underestimates the dengue counts by excluding many asymptomatic and mild dengue cases due to its case definition [37]. Thus, the only national surveillance data likely underrepresents the true magnitude of reported dengue cases. On the other hand, dengue spread is a multidimensional and complex phenomenon, and past studies have outlined that climatic parameters explain 10-30% of this dynamics [51,63,64]. Because of data constraints, it was not possible to incorporate unmeasured confounders, such as variations in virus serotypes, dengue vector population density, urban morphology, infrastructure information, socioeconomic factors, public health interventions, and vector control measures.

While there may have been some constraints in this study, our findings still provide valuable insights into the short-term pattern of dengue occurrence in Dhaka city under current and future climatic conditions. To the best of our knowledge, investigation of the non-linear and lagged association of climatic variables and heatwaves on dengue cases, considering an extended timeline, has not been previously performed. In hyperendemic regions such as Dhaka city, analytical models based on syndromic case surveillance, such as our study, may offer more useful information than vector densities, since DENV transmission can still occur despite low dengue vector population densities due to their repeated feeding patterns [65,66]. Moreover, the findings could also provide valuable information regarding precautionary measures against several other arboviruses such as chikungunya, yellow fever, and Zika, as *Aedes* mosquitoes serve as vectors for these in addition to the DENV.

## Conclusion

This study intends to contribute further to the existing body of knowledge on dengue by providing insights into the exposure-response relationship of climatic variables and dengue cases in Dhaka city. The findings of the study suggest that the maximum temperature range of 31.5-33.2°C demonstrated the highest short-term risk of dengue cases in Dhaka city; however, sustained extreme temperatures (> 35.5°C) could suppress the risk. These quantitative observations can be instrumental in understanding the interaction between vectors and viruses in present and future extreme climatic conditions in dengue-devastated Dhaka city, resulting in a better prediction of disease transmission pathways. Moreover, they would further bolster the efficacy of designing and implementing early warning systems, dengue prevention strategies and vector control measures by policymakers and authorities.

## Data Availability

All relevant data and code are in the GitHub (https://github.com/AhnafShahriyar/Dengue_MaxT_Heatwave_DLNM_Dhaka)

## Ethics statement

The protocol of this study was approved by the Committee for Advanced Studies & Research (CASR), Bangladesh University of Engineering and Technology (BUET). The study used national surveillance data, which neither involved human participants nor collected personally identifiable information.

## CRediT authorship contribution statement

**Ahnaf Shahriyar:** Conceptualization, Data curation, Formal analysis, Investigation, Methodology, Visualization, Writing - Original Draft. **S.M. Manzoor Ahmed Hanifi:** Validation, Writing - Review & Editing. **Sheikh Mokhlesur Rahman:** Conceptualization, Funding Acquisition, Methodology, Resources, Supervision, Validation, Writing - Review & Editing.

## Declaration of generative AI and AI-assisted technologies in the writing process

During the preparation of this work the authors used OpenAI’s ChatGPT and QuillBot AI in order to paraphrase, improve readability and language of the work. After using this tool/service, the authors reviewed and edited the content as needed and take full responsibility for the content of the published article.

## Supporting Information (SI)

**S1 Eq. Calculating Absolute Humidity (AbsHum).**

**S1 Table. Example Calculation for Determining Heatwave Days.** Days with a daily maximum temperature exceeding the 90^th^ percentile threshold of 35.5℃ are shown in bold. Heatwave days are identified when there are at least two consecutive days above this threshold. Heatwaves that start on the Saturday of the previous week and continue into the current week are also included in the count.

**S2 Table. Counts of Weeks with a Total of n Heatwave Days Under Different Heatwave Definitions in Sensitivity Analysis.**

**S1 Fig. Weekly Heatwave Days of Dhaka City (2014-2024) for Different ‘Heatwave Day’ Definitions.** ‘HW_80_2’ denotes number of heatwave days per week where maximum temperature exceeding 80^th^ percentile for 2 consecutive days

**S2 Fig. Time Series of Dengue Admitted Cases of Eight Divisions of Bangladesh (2016-2024).**

**S3 Fig. Weekly Dengue Cases of Dhaka City (2014-2024), Categorized as Outbreak Status.** ‘Small outbreaks’ were classified as weeks with case counts between the 50^th^ and 75^th^ percentiles, medium outbreaks between the 75^th^ and 90^th^ percentiles, and large outbreaks exceeded the 90^th^ percentile

**S4 Fig. Time Series of Daily Climatic Parameters of Dhaka City (2016-2024).**

**S5 Fig. Spearman Correlation Heatmap of Weekly Dengue Infections and Meteorological Data of Dhaka City (2016-2024).**

**S3 Table. Generalized Variance Inflation Factor (GVIF) Values – MaxT Model.**

**S6 Fig. Residual Analysis for DLNM-MaxT Model.** (a) Residual Autocorrelation Function (ACF) in the final MaxT model; (b) Residual Partial Autocorrelation Function (PACF) in the final MaxT model; (c) Residual histogram; (d) Residual vs log of dengue cases per week.

**S4 Table. Sequential Analysis of Deviance for the MaxT Model.**

**S7 Fig. Plot of Exposure–Response Associations at Each Lag Week (0–6) for the MaxT Model.**

**S8 Fig. Sensitivity Analysis Using Varying Degree of Control for Long-Term Trend and Seasonality – MaxT Model.** (a) Natural Cubic Spline of Time (Week), ns(t,df) with 5 df per Year, and (b) Natural Cubic Spline of Time (Week), ns(t,df) with 7 df per Year. All exposure-response curves are computed with reference to mean value of MaxT, 31.5°C. Shaded areas represent 95% confidence intervals.

**S5 Table. Sensitivity Analysis Varying the Degrees of Freedom for the Non-Linear and Lagged Effects in the MaxT Cross-Basis Matrix.**

**S6 Table. Generalized Variance Inflation Factor (GVIF) Values – Heatwave Days (HW) Model.**

**S9 Fig. Residual Analysis for DLNM-Heatwave Model.** (a) Residual Autocorrelation Function (ACF) in the final Heatwave model; (b) Residual Partial Autocorrelation Function (PACF) in the final Heatwave model; (c) Residual histogram; (d) Residual vs log of dengue cases per week.

**S7 Table. Sequential Analysis of Deviance for the HW Model.**

**S10 Fig. Plot of Exposure–Response Associations at Each Lag Week (0–6) for the Heatwave Days Model.**

**S11 Fig. Sensitivity Analysis by Varying Degree of Control for Long-Term Trend and Seasonality in HW Model.** Natural Cubic Spline of Time (Week) with (a) 4 df per year, (b) 5 df per year, and (c) 7 df per year. All exposure-response curves are computed with reference to no heatwave days in the week.

**S12 Fig. Sensitivity Analysis Using Different Heatwave Cross-Basis Specifications.** (a) 2 internal knots at 2 and 4 heatwave days (3 df), (b) 2 internal knots at 2 and 5 heatwave days (3 df), (c) 2 internal knots at 2 and 6 heatwave days (3 df), (d) 2 internal knots at 3 and 5 heatwave days (3 df), (e) 2 internal knots at 3 and 6 heatwave days (3 df), (f) 3 internal knots at 2, 3, and 6 heatwave days (4 df), and (g) 4 internal knots at 2, 3, 4, and 6 heatwave days (5 df). All exposure-response curves are computed with reference to no heatwave days in the week.

**S13 Fig. Sensitivity Analysis Using Alternative Heatwave Definitions.** (a) maximum temperature exceeding the 90^th^ percentile (35.5°C) for two or more consecutive days ‒ original condition; (b) exceeding the 90^th^ percentile for three or more days; (c) exceeding the 80^th^ percentile (34.5°C) for two or more days; (d) exceeding the 80^th^ percentile for three or more days; (e) exceeding the 95^th^ percentile (36.3°C) for two or more days; and (f) exceeding the 95^th^ percentile for three or more days. All exposure-response curves are computed relative to weeks with no heatwave days.

**S14 Fig. Exposure-response curve showing overall cumulative effect of average relative humidity (AH).** RR of reported dengue infections for lag period 0-6 weeks, with reference its mean value, 72.67(%), for (a) Heatwave model and the (b) MaxT model. Shaded areas indicate 95% confidence intervals.

**S15 Fig. Exposure-response curve showing overall cumulative effect of cumulative rainfall (CRF).** RR of reported dengue infections for lag period 0-6 weeks, with reference its mean value, 37.71 mm, for (a) Heatwave model and the (b) MaxT model. Shaded areas indicate 95% confidence intervals.

## Acknowledgement

The authors would like to acknowledge the financial and academic support of Committee for Advanced Studies and Research (CASR), Bangladesh University of Engineering and Technology (BUET) for undertaking this research. The authors would further like to acknowledge Bangladesh Meteorological Department (BMD) and Directorate General of Health Services (DGHS), Bangladesh for providing climate and dengue cases data.

